# Effects of Empagliflozin on Right Ventricular Function in Heart Failure with Reduced Ejection Fraction

**DOI:** 10.1101/2025.05.30.25328686

**Authors:** Massar Omar, Jesper Jensen, Peter H. Frederiksen, Mulham Ali, Caroline Kistorp, Lars Videbæk, Mikael Kjær Poulsen, Christian D. Tuxen, Barry A. Borlaug, Sören Möller, Finn Gustafsson, Lars Køber, Morten Schou, Jacob Eifer Møller

## Abstract

**Objectives:** To investigate the effect of empagliflozin on right ventricular (RV) function in patients with HFrEF.

**Background:** Sodium-glucose cotransporter-2 (SGLT2) inhibitors improve outcomes and reverse left ventricle (LV) remodeling in heart failure with reduced ejection fraction (HFrEF). The impact on RV function remains uncertain.

**Methods:** The Empire HF trial was a randomized, double-blind trial of 190 participants with HFrEF (LVEF ≤40%) randomized (1:1) to empagliflozin (10mg/daily) or matching placebo for 12 weeks. The primary endpoint was changes in RV free wall strain (RVFWS) across the whole cohort. RVFWS was also stratified into tertiles based on baseline RVFWS. Secondary endpoints included changes in TAPSE and RV S’ velocity.

**Results:** Baseline characteristics were balanced between the groups (mean age of 64±11 years, 86% male, mean LVEF 29±8%, 79% in NYHA class II). The overall mean RVFWS was −16.7±6.1%, with the empagliflozin group at −16.4±6.2% and the placebo group at −16.9±5.9%. No differences were observed in RVFWS between the groups. When stratified by baseline RVFWS into tertiles, patients in the lowest tertile demonstrated a significant improvement with empagliflozin (treatment effect: −2.9% [95% CI: −5.0 to −0.3]; P=0.027), independent of LVEF, plasma volume, or weight loss changes. No significant treatment effect was observed in the middle or highest tertiles of RVFWS, nor in the overall or lowest tertiles of TAPSE or RV S’.

**Conclusions:** Empagliflozin exerted no overall effect on RV function in stable HFrEF patients, but significantly improved RVFWS in patients in the lowest tertile of RVFWS after 12 weeks of treatment.

**Trial Registration:** ClinicalTrials.gov, Unique Identifier: NCT03198585

## Introduction

Dysfunction of the right ventricle (RV) is common in heart failure (HF) with reduced ejection fraction (HFrEF), occurring in 35-50% of patients (1). RV dysfunction is associated with progression and prognosis in HF patients (2), with earlier studies showing that each 1% worsening in RV free wall strain (RVFWS) was independently associated with a 5% increased risk of all-cause mortality (3).

Sodium-Glucose Transporter-2 (SGLT2) inhibitors have emerged as a potent HFrEF therapy, improving clinical outcomes and quality of life in patients with HF irrespective of left ventricular ejection fraction (LVEF) (4–6). The exact mechanisms underlying the beneficial effects remain partially elucidated. The benefits may be related to related to improved hemodynamics including, but not restricted to, reduced preload, reverse left ventricular (LV) remodeling, and metabolic changes (7–9).

Knowledge regarding the effect of SGLT2 inhibitors on RV function remains remarkably limited. A small, single-blinded study conducted in 36 patients with HFrEF, along with two observational studies, found that empagliflozin significantly improved the systolic and global RV function assessed by RVFWS and tricuspid annular plane systolic excursion (TAPSE) after 12 and 24 weeks of treatment, respectively (10–12).

However, observational studies are subject to bias and cannot provide definite proof of the effect of an intervention. To distinguish treatment effects from the natural progression of the disease, a control group is required. Therefore, the aim of this post-hoc analysis of the Empagliflozin in heart failure patients with reduced ejection fraction (Empire HF) trial was to investigate the impact of empagliflozin on RV function in 190 patients with HFrEF after 12 weeks of treatment compared to placebo (13).

## Methods

### Study design and ethics

This is a post-hoc analysis from the Empire HF trial which has been previously published (13,14). The trial was an investigator-initiated, multicenter, randomized, double-blind, placebo-controlled clinical trial, randomly assigning 190 patients with HFrEF (1:1) to empagliflozin 10 mg once daily or matching placebo for 12 weeks. At baseline, patients underwent a comprehensive clinical examination, transthoracic echocardiography, and blood tests, which were repeated at 12-weeks follow-up.

The trial was conducted in accordance with Good Clinical Practice and the Declaration of Helsinki. All participants signed informed consent before inclusion. The Empire-HF trial is registered in ClinicalTrials.gov, NCT03198585.

### Study Participants

Stable HFrEF patients aged ≥18 years, on optimal HF therapy in accordance with European and national guidelines, with a LVEF ≤40%, and with a New York Heart Association (NYHA) class I-III, were eligible for this study. The full list of inclusion and exclusion criteria is presented in the appendix (*Appendix, section C)*

### Echocardiography

Transthoracic echocardiography was performed on a Vivid e9 ultrasound system (General Electric, Horten, Norway), and stored digitally for offline analysis using Echopac (version 203, General Electric). Echocardiographic measurements were not pre-specified in the statistical analysis plan and were decided after the end of the trial (14), but before un-blinding of the study. Analyses were performed in the intention-to-treat population, including all randomized patients with complete data, analyzed in a random order and blinded to treatment allocation.

The function of the RV was assessed by measuring RVFWS, TAPSE, and RV S’ wave in accordance with current guidelines (15). TAPSE was measured in the Apical 4 Chamber (A4C) - view using the M-mode cursor over the lateral tricuspid annulus and measuring a straight line from top to bottom of the wave, which calculates the systolic displacement of the tricuspid annulus, recorded in cm. RV S’ wave was measured using tissue Doppler imaging (TDI) from the A4C view by placing the Doppler cursor on the lateral tricuspid annulus to record the peak systolic velocity (S’ wave) in cm/s, representing RV longitudinal systolic function. The maximal value was chosen for analysis. RVFWS was measured using Q-analysis of the Echopac software from the apical four-chamber view. The region of interest was set to cover the tricuspid insertion, from the lateral RV free wall to the RV apex. RVFWS is calculated as the percentage of systolic shortening of the RV free wall from base to apex (15), and is reported as a negative value.

LV volumes and LVEF were assessed using the biplane method of disks (Simpsonśs Biplane). A detailed description of the echocardiography protocol is provided in the Appendix *(Appendix, section D)*.

### Efficacy measures

The primary efficacy measure was the between-group difference in the change in RVFWS from baseline to 12 weeks follow-up. Secondary exploratory measures included the between-group differences in the changes of TAPSE and RV S’ velocity.

### Statistical analysis

No specific sample size estimation was performed for this post-hoc analysis, and study population size was the sample size of the main Empire HF trial (14). The primary statistical analysis was based on all included patients with available RVFWS measurement based on the per protocol principle. We replicated the analyses with the full study cohort on available data for TAPSE and RV S’, and findings remained consistent with the intention-to-treat in a sensitivity analysis *(Appendix, Table 3)*.

Baseline characteristics and echocardiographic measurements were reported as means and standard deviations (SD) for normally distributed variables, numbers (%) for categorical variables, and medians with interquartile ranges (IQR) for non-normally distributed variables.

Patients were stratified into subgroups based on tertiles of baseline values of RVFWS. The subgroups values were derived from the distribution of RVFWS at baseline in the study population, as the existing literature does not provide a consensus on the appropriate diagnostic and prognostic cut-off values for patients with HFrEF (16). Furthermore, to visualize and delineate the associations of treatment effect on RVFWS, we further tested the non-linear relationship between empagliflozin versus placebo on RVFWS (as a continuous variable) using restricted cubic spline curves. The between group difference was based on a per protocol analysis, adjusted for age, sex, diabetes, atrial fibrillation, and coronary artery bypass grafting (CABG).

Subgroup analyses were interpreted with mean change and 95% confidence intervals (CI) as the change between the groups by the specified subgroups. The *P*-value denotes the two-way interaction between subgroups.

All statistical tests were performed for a two-sided 0.05 alpha-level of significance and reported with the corresponding 95% CI. Statistical analyses were conducted using STATA statistical software, version 17 (Stata Crop, College. Station, Texas, USA).

## Results

### Baseline characteristics

The groups did not differ with respect to baseline characteristics (*Table 1*). The mean age was 64 years, 138 (86%) were male, 126 (79%) reported a NYHA class II, and a high proportion on optimal medical treatment for HFrEF. There were no clinically meaningful between-group differences in concomitant HF therapy, including diuretic dosages at baseline (*Table 1*), and no significant changes between groups during the study period. Both baseline and 12-week follow-up echocardiographic measurements of RVFWS were available for 82 patients in the empagliflozin group and 78 in the placebo group, and these patients were included in the primary intention-to-treat analysis *(Appendix, Figure 1)*. All patients, except one, had medication compliance greater than 90% *(Appendix, Figure 1)*.

**Figure 1:**
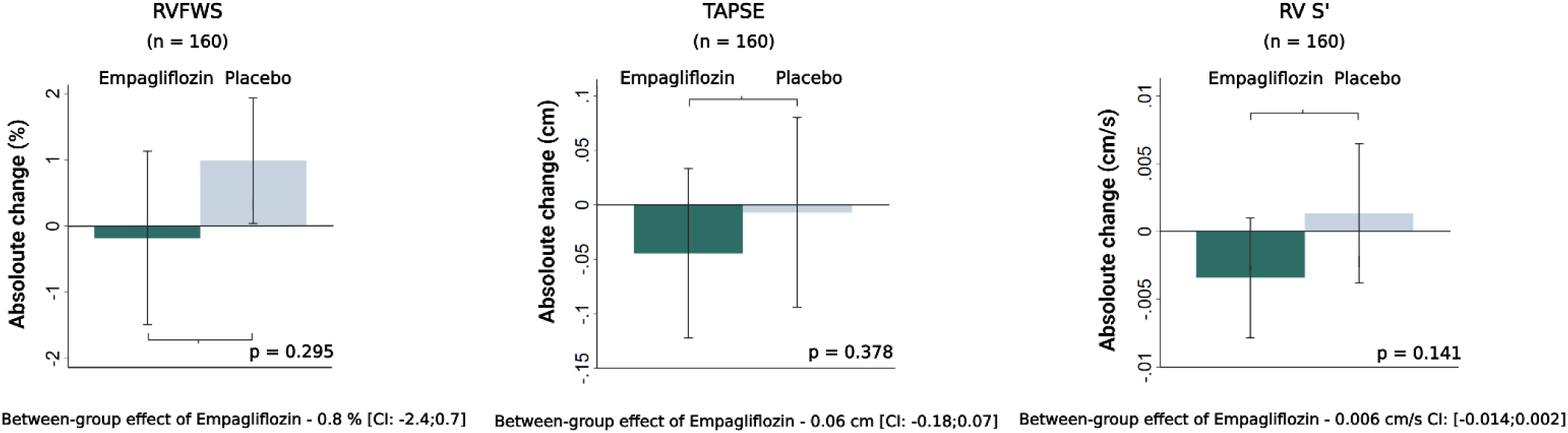
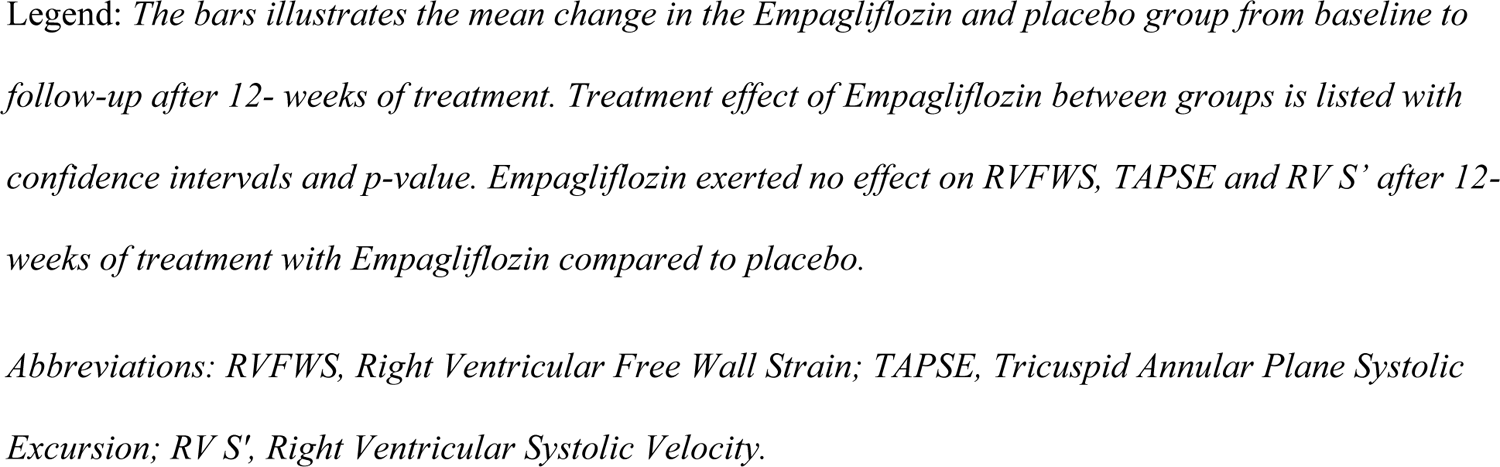
Effects on Right Ventricular parameters in the total population

**Table 1:**
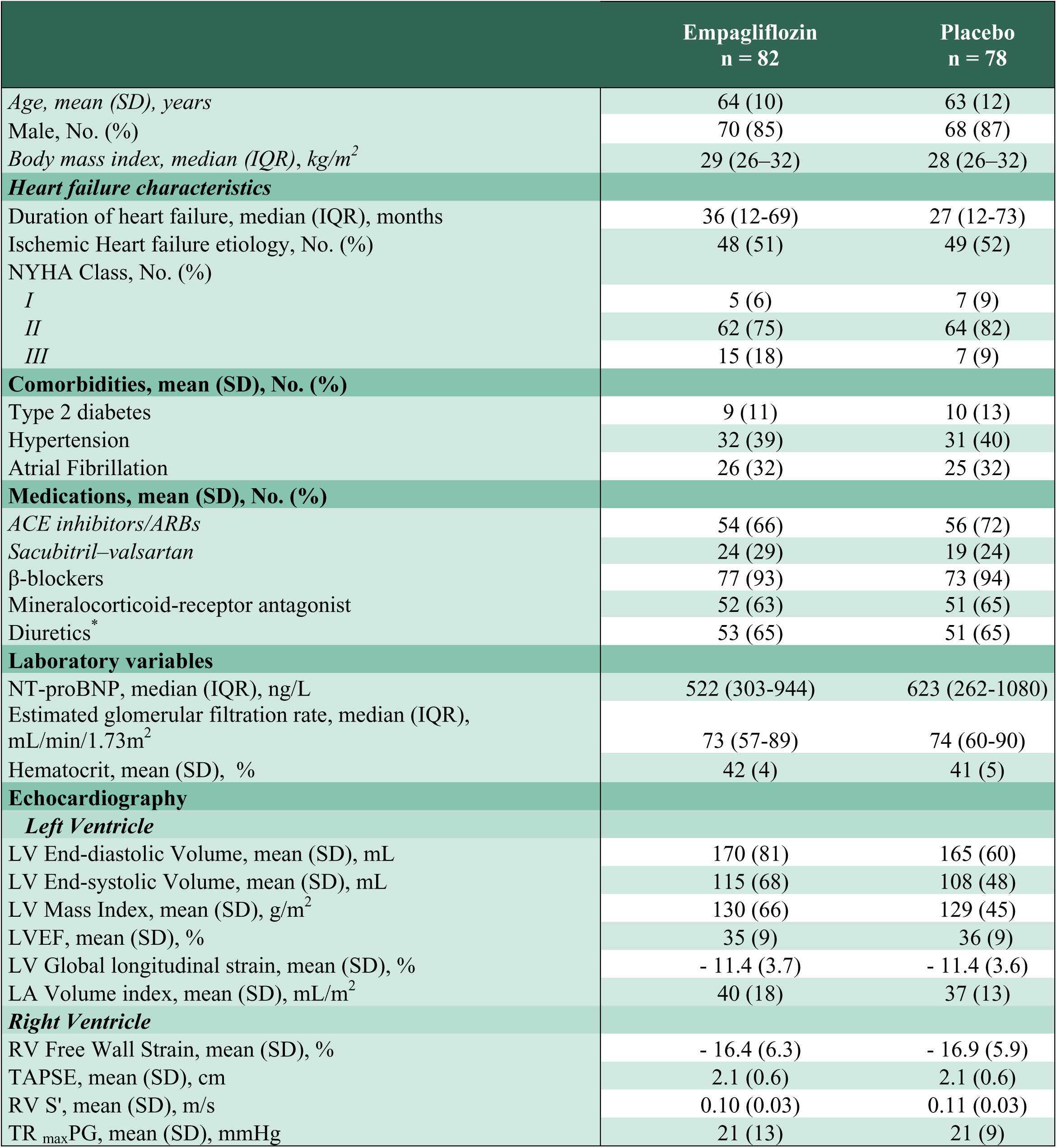
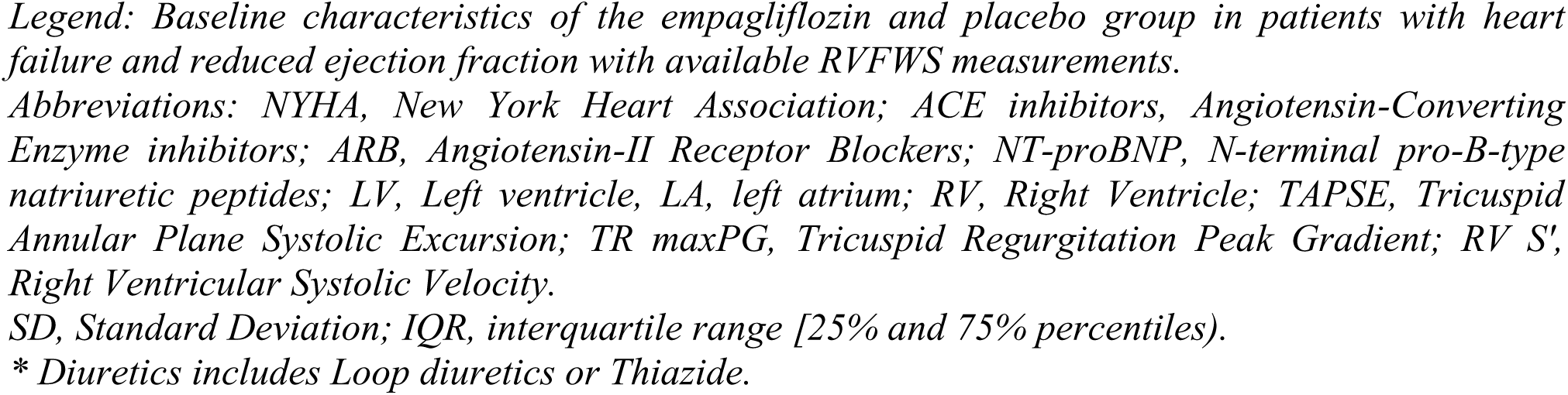
Baseline Characteristics.

Baseline characteristics of the tertiles of RVFWS are presented in Table 2. The baseline means of RVFWS were −23.3% (SD, 4.4), −16.0% (SD, 1.4) and −10.5% (SD, 2.6) for the upper, middle and lower tertiles, respectively. Patients in the lower tertile of RVFWS had a longer duration of HFrEF, lower LVEF, higher body weight, and a greater prevalence of diabetes and atrial fibrillation, with a higher proportion (77%) of them on diuretics, compared to 65% in the entire cohort. Additionally, they had the highest LV end-diastolic and end-systolic volumes, LV mass index, left atrial volume index, RV S’, and the lowest LVEF compared to middle and upper tertile groups. At baseline, there were no significant differences between subgroups in use of guideline-directed medical therapy (GDMT) including diuretics or in estimated glomerular filtration rate (*Table 2)*.

**Table 2:**
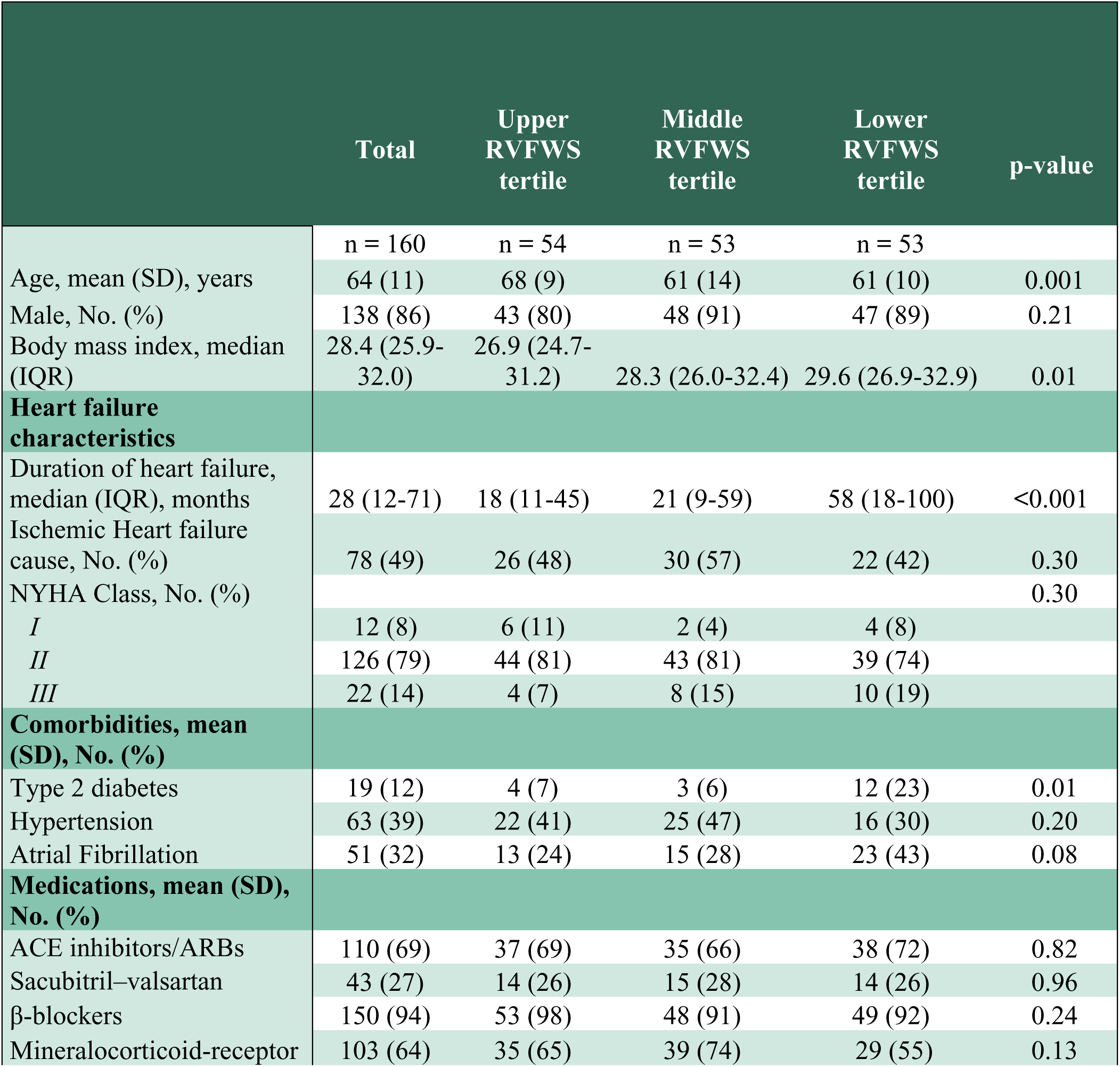
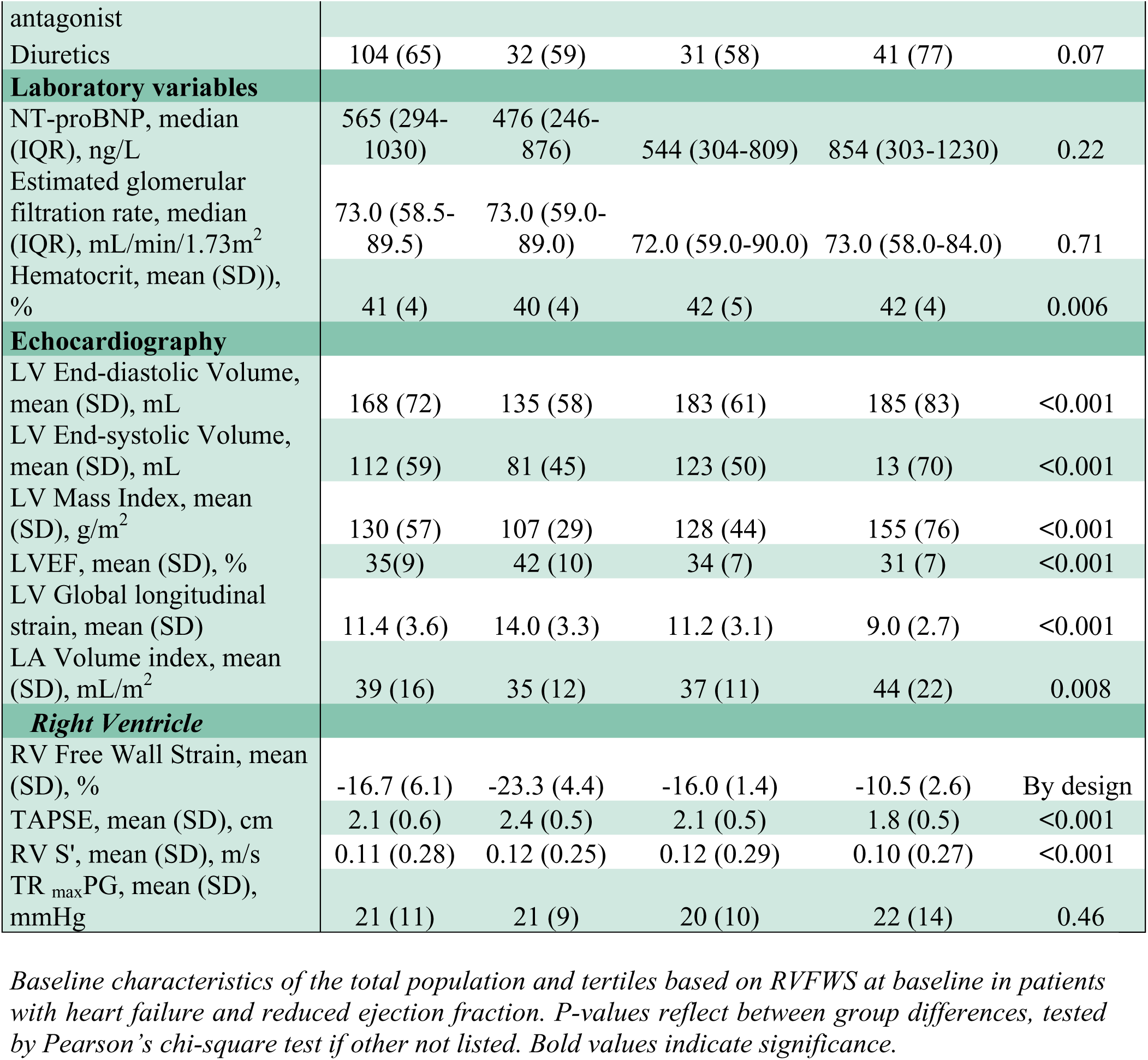
Baseline Characteristics Across Subgroups.

### Primary efficacy measure

At baseline, participants had an overall RVFWS mean of −16.7% (SD 6.1), with the empagliflozin group at −16.4% (SD 6.2) and the placebo group at −16.9% (SD 5.9). The overall between-group treatment effect after 12 weeks was not statistically significant (adjusted treatment effect: −0.8% [95% CI: −2.4 to 0.7]; *P =* 0.30) (*Table 3, Figure 1*).

Baseline means of the RVFWS tertiles were as follows: for the empagliflozin group, the upper tertile was −23.4% (SD 3.9), middle tertile −15.9% (SD 1.6), and lower tertile −9.7% (SD 2.4); for the placebo group, −23.2% (SD 4.9), −16.1% (SD 1.2), and −11.3% (SD 2.6), respectively (*Table 3*). In patients with the lowest RVFWS, empagliflozin treatment was associated with a significant improvement in RVFWS from baseline to follow-up compared to the placebo group (adjusted treatment effect: −2.9% [95% CI:-5.0 to −0.3]; P=0.03) (*Table 3, Figure 2*); There were no differences between empagliflozin and placebo in the upper or middle RVFWS tertiles (*Table 2, Figure 1*).

**Figure 2:**
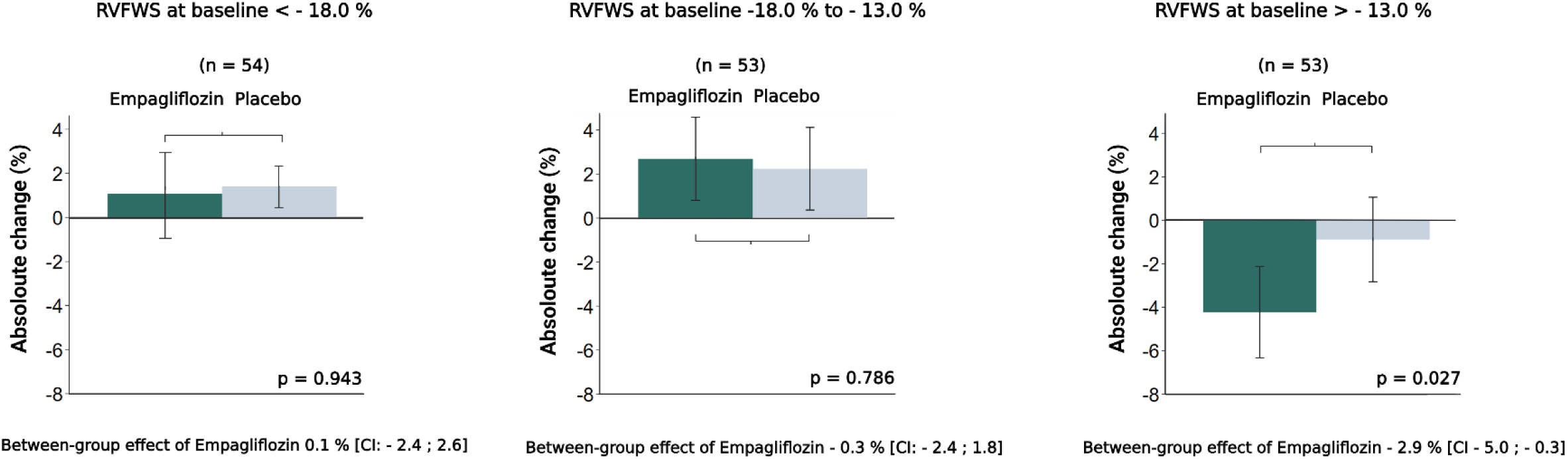
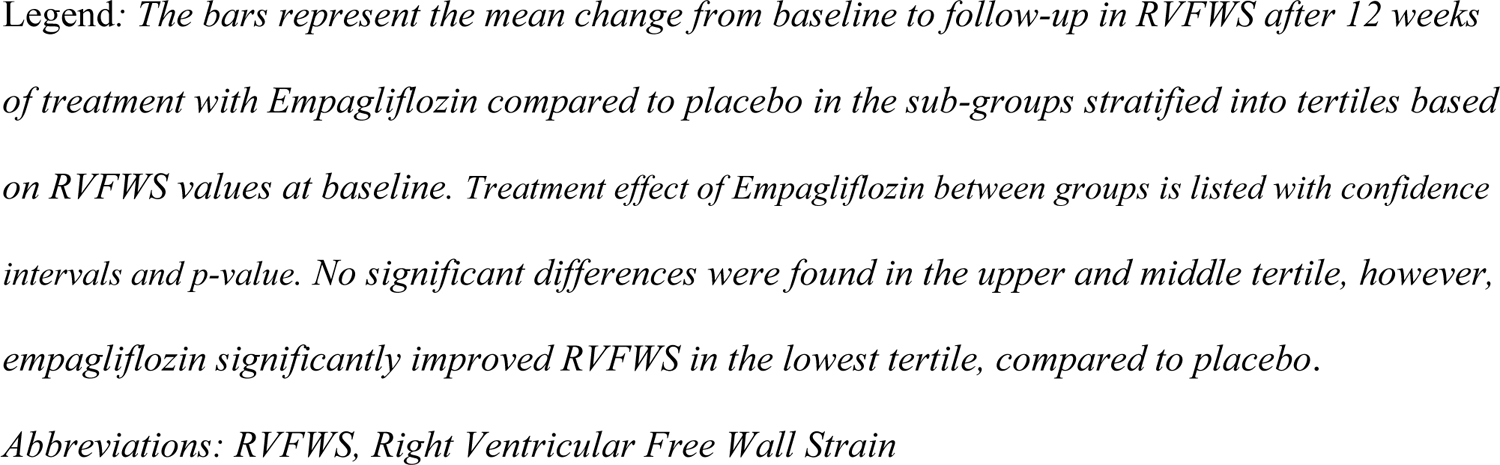
Effect on Right Ventricular Free Wall Strain across Subgroups

**Table 3:**
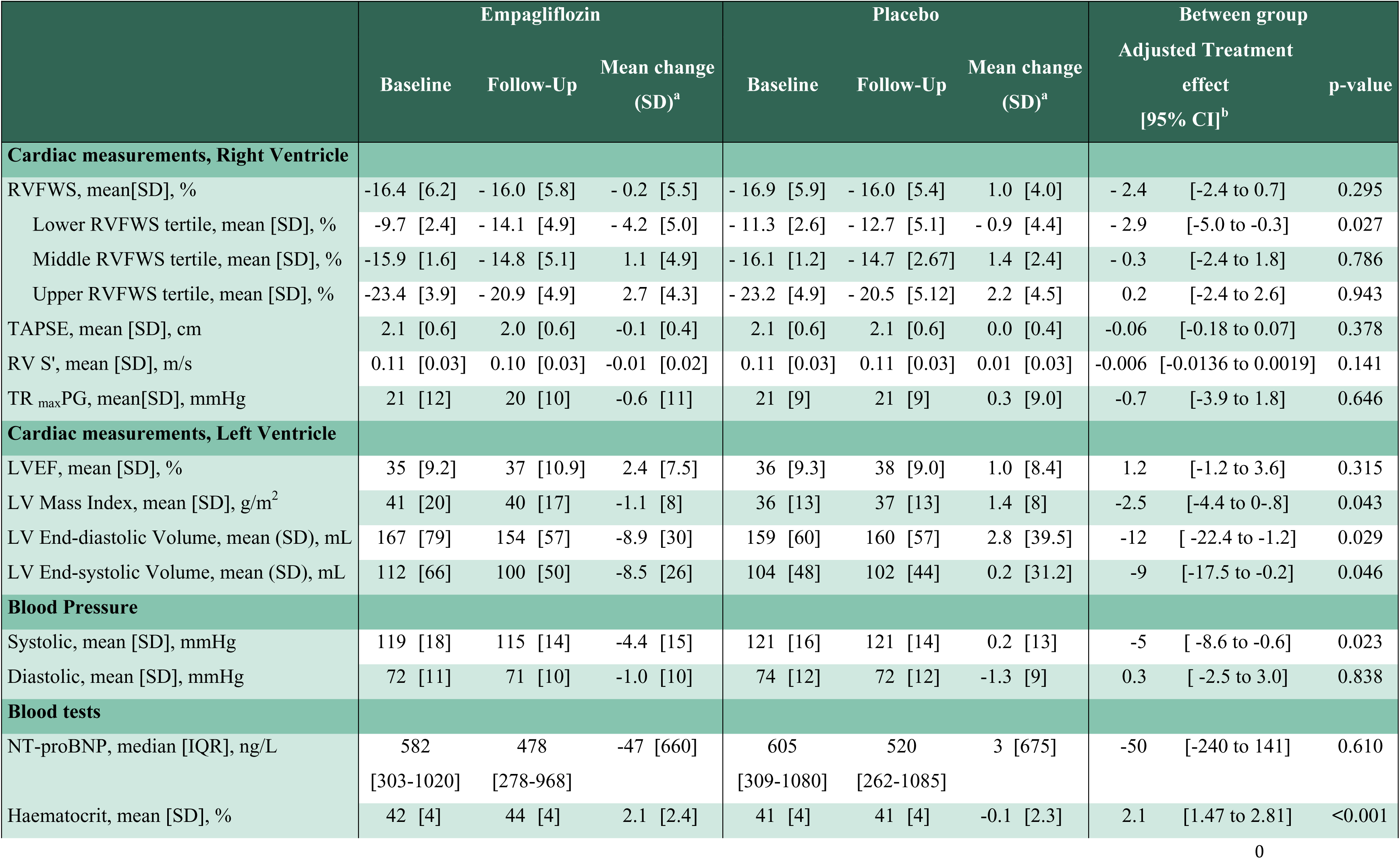
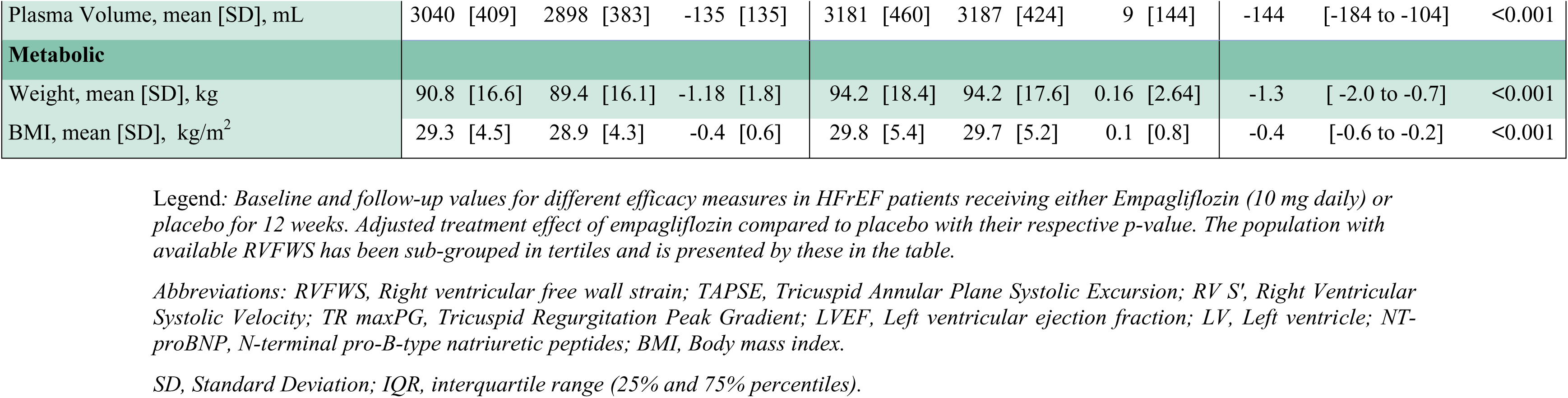
Efficacy Measures.

|  | Empagliflozin |  |  | Placebo |  |  | Between group |  |
| --- | --- | --- | --- | --- | --- | --- | --- | --- |
|  | Baseline | Follow-Up | Mean change<br>(SD)<sup>a</sup> | Baseline | Follow-Up | Mean change<br>(SD)<sup>a</sup> | Adjusted Treatment<br>effect<br>[95% CI]<sup>b</sup> | p-value |
| Cardiac measurements, Right Ventricle |  |  |  |  |  |  |  |  |
| RVFWS, mean[SD], % | -16.4 [6.2] | - 16.0 [5.8] | - 0.2 [5.5] | - 16.9 [5.9] | - 16.0 [5.4] | 1.0 [4.0] | - 2.4 [-2.4 to 0.7] | 0.295 |
| Lower RVFWS tertile, mean [SD], % | -9.7 [2.4] | - 14.1 [4.9] | - 4.2 [5.0] | - 11.3 [2.6] | - 12.7 [5.1] | - 0.9 [4.4] | - 2.9 [-5.0 to -0.3] | 0.027 |
| Middle RVFWS tertile, mean [SD], % | -15.9 [1.6] | - 14.8 [5.1] | 1.1 [4.9] | - 16.1 [1.2] | - 14.7 [2.67] | 1.4 [2.4] | - 0.3 [-2.4 to 1.8] | 0.786 |
| Upper RVFWS tertile, mean [SD], % | -23.4 [3.9] | - 20.9 [4.9] | 2.7 [4.3] | - 23.2 [4.9] | - 20.5 [5.12] | 2.2 [4.5] | 0.2 [-2.4 to 2.6] | 0.943 |
| TAPSE, mean [SD], cm | 2.1 [0.6] | 2.0 [0.6] | -0.1 [0.4] | 2.1 [0.6] | 2.1 [0.6] | 0.0 [0.4] | -0.06 [-0.18 to 0.07] | 0.378 |
| RV S', mean [SD], m/s | 0.11 [0.03] | 0.10 [0.03] | -0.01 [0.02] | 0.11 [0.03] | 0.11 [0.03] | 0.01 [0.03] | -0.006 [-0.0136 to 0.0019] | 0.141 |
| TR<sub>max</sub>PG, mean[SD], mmHg | 21 [12] | 20 [10] | -0.6 [11] | 21 [9] | 21 [9] | 0.3 [9.0] | -0.7 [-3.9 to 1.8] | 0.646 |
| Cardiac measurements, Left Ventricle |  |  |  |  |  |  |  |  |
| LVEF, mean [SD], % | 35 [9.2] | 37 [10.9] | 2.4 [7.5] | 36 [9.3] | 38 [9.0] | 1.0 [8.4] | 1.2 [-1.2 to 3.6] | 0.315 |
| LV Mass Index, mean [SD], g/m<sup>2</sup> | 41 [20] | 40 [17] | -1.1 [8] | 36 [13] | 37 [13] | 1.4 [8] | -2.5 [-4.4 to 0-.8] | 0.043 |
| LV End-diastolic Volume, mean (SD), mL | 167 [79] | 154 [57] | -8.9 [30] | 159 [60] | 160 [57] | 2.8 [39.5] | -12 [ -22.4 to -1.2] | 0.029 |
| LV End-systolic Volume, mean (SD), mL | 112 [66] | 100 [50] | -8.5 [26] | 104 [48] | 102 [44] | 0.2 [31.2] | -9 [-17.5 to -0.2] | 0.046 |
| Blood Pressure |  |  |  |  |  |  |  |  |
| Systolic, mean [SD], mmHg | 119 [18] | 115 [14] | -4.4 [15] | 121 [16] | 121 [14] | 0.2 [13] | -5 [ -8.6 to -0.6] | 0.023 |
| Diastolic, mean [SD], mmHg | 72 [11] | 71 [10] | -1.0 [10] | 74 [12] | 72 [12] | -1.3 [9] | 0.3 [ -2.5 to 3.0] | 0.838 |
| Blood tests |  |  |  |  |  |  |  |  |
| NT-proBNP, median [IQR], ng/L | 582<br>[303-1020] | 478<br>[278-968] | -47 [660] | 605<br>[309-1080] | 520<br>[262-1085] | 3 [675] | -50 [-240 to 141] | 0.610 |
| Haematocrit, mean [SD], % | 42 [4] | 44 [4] | 2.1 [2.4] | 41 [4] | 41 [4] | -0.1 [2.3] | 2.1 [1.47 to 2.81] | <0.001 |
| Plasma Volume, mean [SD], mL | 3040 [409] | 2898 [383] | -135 [135] | 3181 [460] | 3187 [424] | 9 [144] | -144 [-184 to -104] | <0.001 |
| **Metabolic** |  |  |  |  |  |  |  |  |
| Weight, mean [SD], kg | 90.8 [16.6] | 89.4 [16.1] | -1.18 [1.8] | 94.2 [18.4] | 94.2 [17.6] | 0.16 [2.64] | -1.3 [-2.0 to -0.7] | <0.001 |
| BMI, mean [SD], kg/m<sup>2</sup> | 29.3 [4.5] | 28.9 [4.3] | -0.4 [0.6] | 29.8 [5.4] | 29.7 [5.2] | 0.1 [0.8] | -0.4 [-0.6 to -0.2] | <0.001 |

Legend: *Baseline and follow-up values for different efficacy measures in HFrEF patients receiving either Empagliflozin (10 mg daily) or*
*placebo for 12 weeks. Adjusted treatment effect of empagliflozin compared to placebo with their respective p-value. The population with*
*available RVFWS has been sub-grouped in tertiles and is presented by these in the table.*
*Abbreviations: RVFWS, Right ventricular free wall strain; TAPSE, Tricuspid Annular Plane Systolic Excursion; RV S', Right Ventricular*
*Systolic Velocity; TR maxPG, Tricuspid Regurgitation Peak Gradient; LVEF, Left ventricular ejection fraction; LV, Left ventricle; NT-*
*proBNP, N-terminal pro-B-type natriuretic peptides; BMI, Body mass index.*
*SD, Standard Deviation; IQR, interquartile range (25% and 75% percentiles).*

As depicted in Figure 3, patients with the lowest baseline RVFWS values experienced the greatest improvement with empagliflozin treatment, corroborating the tertile stratification analysis.

**Figure 3:**
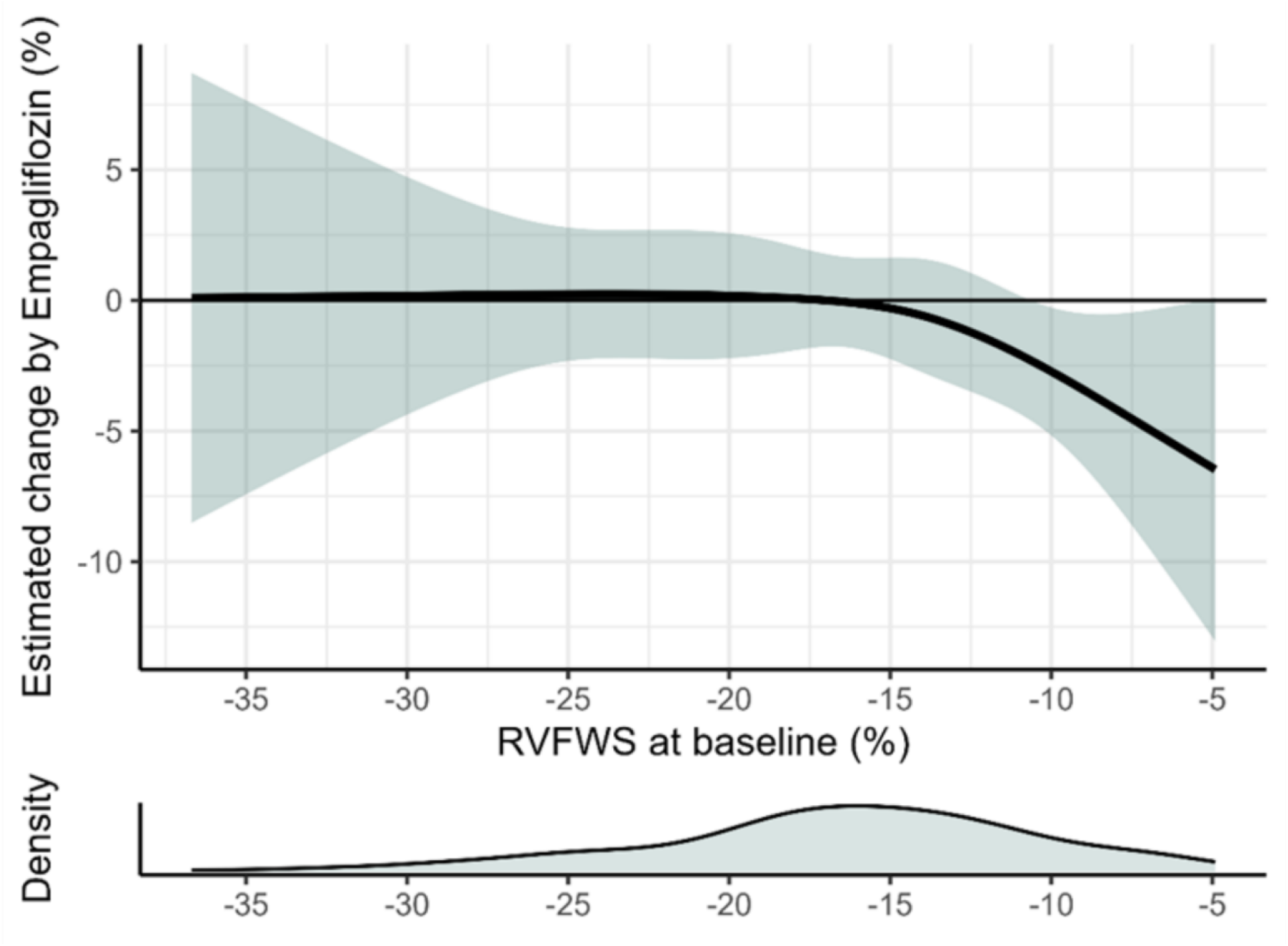
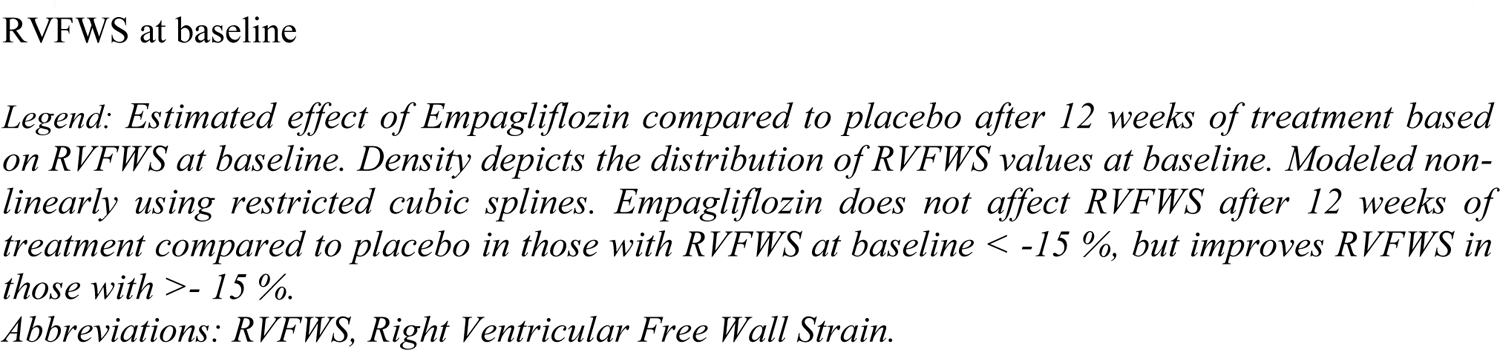
Estimated change in RVFWS by 12 weeks of treatment with Empagliflozin relatively to RVFWS at baseline

The improvement in RVFWS was not associated with weight loss, plasma volume, systolic blood pressure, or hematocrit (P > 0.05), nor were any significant correlations with changes in LV and RV function or structure found (*P > 0.05 for all*) (*Appendix, Table 2*).

### Secondary efficacy measures and correlations

Compared to placebo, empagliflozin did not have a significant impact on TAPSE (adjusted treatment effect: −0.06 cm [95% CI: −0.18 to 0.07]; P=0.78) or RV S’ (adjusted treatment effect: −0.006 cm/s [95%: −0.014 to 0.002]; P=0.14), on the overall population *(Table 3, Figure 1).* These findings were consistent when patients were stratified into baselines tertiles of TASPE and RV S’ *(Appendix, Table 3)*.

## Discussion

To our knowledge, this is the first study to report the effect of SGLT2 inhibition on RV function in a double-blind, randomized, controlled trial including a sizable cohort of stable, euvolemic patients with HFrEF. The study shows that empagliflozin did not yield a discernible effect on RV function in the overall study population compared to placebo after 12 weeks of treatment, but significantly improved RVFWS in patients with the lowest RVFWS at baseline. No significant changes were seen in TAPSE or the RV S’ velocity.

A smaller single-blinded, prospective study of 36 patients with HFrEF showed that empagliflozin significantly improved RVFWS after 12 weeks (baseline RVFWS −16.4%) (10). Additionally, two observational studies demonstrated improvements in RV systolic function and pulmonary arterial stiffness, with correlations to improved NYHA classification and quality of life (11,12).

In contrast, despite having a similar baseline RVFWS, our study did not find a significant treatment effect in the overall cohort. This discrepancy may be attributed to differences in patient populations. The participants in our study were well-managed, euvolemic, and exhibited only mild symptoms, which may explain why significant improvements in RVFWS were observed exclusively in individuals within the lowest baseline RVFWS tertile. This differs from the three previous studies (11–13), where patients were more decompensated, had larger cardiac chambers, and exhibited greater RV impairment. Conversely, stable HFrEF patients with absent or mild RV dysfunction may not derive a detectable benefit from empagliflozin on RV function, underscoring the potential for patient selection to influence treatment responses. Although the precise mechanisms behind this benefit remain unclear.

We selected RVFWS as the primary efficacy measure because data have shown that it is a better predictor of outcomes than RV global longitudinal strain, purportedly due to being less influenced by LV longitudinal dysfunction (38). Additionally, RVFWS serves not only as a prognostic parameter but also as a marker capable of detecting subtle deterioration in RV systolic function, even when TAPSE and RV S’ wave remain preserved, as observed in our cohort (17–19).

### Diuretics effect

It has been proposed that one of the main mechanisms by which SGLT2 inhibitors exerts their cardio-protective effects is a reduction in preload, primarily due to the diuretic and natriuretic effects (20).

We observed an increase in hematocrit and a decrease in LV end-diastolic volume from baseline to 12 weeks with empagliflozin compared with placebo, consistent with existing studies in the literature (4,6–8). However, the change in RVFWS observed in the subgroup with lowest baseline RVWFS in the current study was not linked to alterations in hematocrit, plasma volume, weight loss, LV structure and function, and as such the observed effects are likely not explained by the diuretic effect of empagliflozin (21).

We did not measure or estimate pulmonary artery pressures, however, a proportion of the study participants (n=70), underwent right-heart catheterization, where we previously found no changes in pulmonary artery pressure or pulmonary vasculature during rest and exercise but a modest decrease in LV filling pressure in the empagliflozin group compared to placebo (20). The pronounced changes in fluid status are the most notable effect of SGLT2 inhibitors, especially in short-term trials. While our findings may be driven by enhanced diuretic efficiency, other mechanisms contributing to the beneficial effects cannot be excluded.

### The “*Thrifty Substrate*” hypothesis

The “*Thrifty Substrate*” hypothesis has been proposed to be part of the SGLT2 inhibitors cardiovascular benefits (27). It has been postulated that shifting fuel utilization from less energy-efficient lipids and glucose toward ketone bodies can enhance myocardial fuel metabolism, myocardial contractility, and cardiac efficiency. Numerous studies have demonstrated that SGLT2 inhibitor cause an increase in ketone bodies in patients with T2D (29,30) and HFrEF (31,32). Interestingly, a recent experimental study demonstrated that increased ketosis in a rodent model, improves RV function (33). Since ketogenesis was not directly measured in our study, a direct link between ketogenesis and improved RV function cannot be established, but the utilization of ketone bodies may additionally contribute to empagliflozin’s improvement of RV function. Dedicated studies specifically testing the effects of SGLT2-induced ketogenesis on RV function, in both clinical human trials and RV remodeling capabilities, are warranted.

### Guideline directed medical therapy and right ventricle

The RV and its impact in HFrEF has generally been neglected by clinicians and researchers. GDMT for HF was almost exclusively developed to target LV dysfunction, resulting in limited knowledge about its effects on RV (dys)function. The question remains whether RV dysfunction *per se* is a therapeutic target or merely may benefit from treatment secondary to improved LV function by improving RV pre-or afterload or as a consequence of ventricular interdependence. It has been found that sacubitril/valsartan improves RV performance and pulmonary hypertension in patients with HFrEF; an effect that was not solely correlated with the reverse remodeling of the LV (34,35). The overall reduced risk of HF events in patients with HFrEF treated with SGLT2 inhibitor, was not diminished in intensively treated patients receiving sacubitril/valsartan (36). As previously reported, we showed that empagliflozin treatment significantly reduced cardiac volumes in patients with HFrEF, and the treatment effect was similar in those receiving sacubitril/valsartan at baseline (37). This present study did not show any correlation between the reverse LV remodeling and the improved RV function, and no significant difference in the distribution of patients receiving sacubitril/valsartan or any other HF GDMT across the tertiles, supporting the beneficial effect beyond the ventricular interdependence of the LV improvement.

The significant improvement in RVFWS observed in the lowest tertile raises the question of regression toward the mean. Patients in this group likely had more impaired RV function at baseline, making spontaneous variation or regression toward the mean a potential contributor to the observed changes. However, the absence of similar improvements in the middle or highest tertiles of RVFWS, as well as in TAPSE or RV S’, suggests that the findings may still reflect a specific treatment effect of empagliflozin in those with more severe RV dysfunction.

### Limitations

The results of this sub-study should be regarded as hypothesis-generating. While the widely used parameter in the clinical setting is TAPSE, RVFWS was chosen as the primary measurement in our study, as RVFWS has been demonstrated to be superior and a more reliable echocardiographic measure compared to TAPSE (3,38), and to correlate better with RV ejection fraction in cardiac magnetic resonance (39). We used echocardiography, which is less accurate and reproducible than cardiac magnetic resonance but remains the most widely utilized method. Despite its limitations, our study showed a consistent effect on RV function in the subgroup analysis.

The HFrEF patients included in this study were relatively young, euvolemic, well-compensated, and the majority were in the NYHA class II. The applicability of our findings to HFrEF patients with more advanced disease remains uncertain and requires further investigation. Moreover, considering the short treatment duration of 12 weeks, it remains unclear whether a more pronounced effect of empagliflozin would have been observed over a longer treatment period.

## Conclusion

This substudy demonstrated that empagliflozin had no significant effect on RV function in the overall cohort of HFrEF patients but significantly improved RVFWS in the lowest tertile, with no changes in the other tertiles after 12 weeks of treatment. These benefits were independent of loading conditions, weight, and LVEF improvement. Further studies are warranted to determine the clinical significance in HFrEF patients

## Clinical Perspective

In euvolemic, stable patients with chronic HFrEF, empagliflozin exerted no overall effect on RV function, but significantly improved RVFWS in patients with the lowest tertile of RVFWS after 12 weeks treatment. The benefits were unrelated to loading conditions, weight, or improvement of LVEF.

## Translational Outlook

Empagliflozin treatment may affect RV function in individuals with the lowest RVFWS values, potentially contributing to the cardioprotective properties of SGLT2 inhibitors. Further research is needed to clarify the mechanisms by which SGLT2 inhibitors improve RV function in patients with HFrEF.

## Role of the Funder/Sponsor

This funding source had no role in the design of the study, its execution, analysis, interpretation of the data, or decision to submit results. There was no interference or financial support from the manufacturer of empagliflozin.

This work was supported by the Danish Heart Foundation (grant numbers 17-R116-A7714-22076, 18-R124-A8573-22107); Steno Diabetes Center Odense, Denmark (grant number 3363); A.P. Møller Foundation for the Advancement of Medical Science (grant numbers 17-L-0339, 17-L-0002); The Research Council at Herlev and Gentofte University Hospital, Denmark (institutional research grant); The Research and Innovation Foundation of the Department of Cardiology (FUHAS, formerly FUKAP), Herlev and Gentofte University Hospital, Denmark (institutional research grant); The Capital Region of Denmark (grant number A6058).

## Abbreviations

HF: Heart Failure
HFrEF: Heart Failure with Reduced Ejection Fraction
LV: Left Ventricle
LVEF: Left ventricular Ejection Fraction
RV: Right Ventricle
RVFWS: Right Ventricular Free Wall Strain
RV S’: Right ventricular peak systolic tissue velocity
SGLT2: Sodium-Glucose cotransporter-2
TAPSE: Tricuspid Annular Plane Systolic Excursion
T2DM: Type 2 Diabetes Mellitus

## Data Availability

The data that support the findings of this study are available from the corresponding author upon reasonable request.

## Acknowledgements

The authors extend their appreciation to all patients for their participation in the trial. We also gratefully acknowledge the dedicated work of all healthcare staff at Odense University Hospital and Herlev-Gentofte Hospital in the trial.

## Central Illustration

Impact of Empagliflozin on Right Ventricular function in Heart Failure with Reduced Ejection Fraction

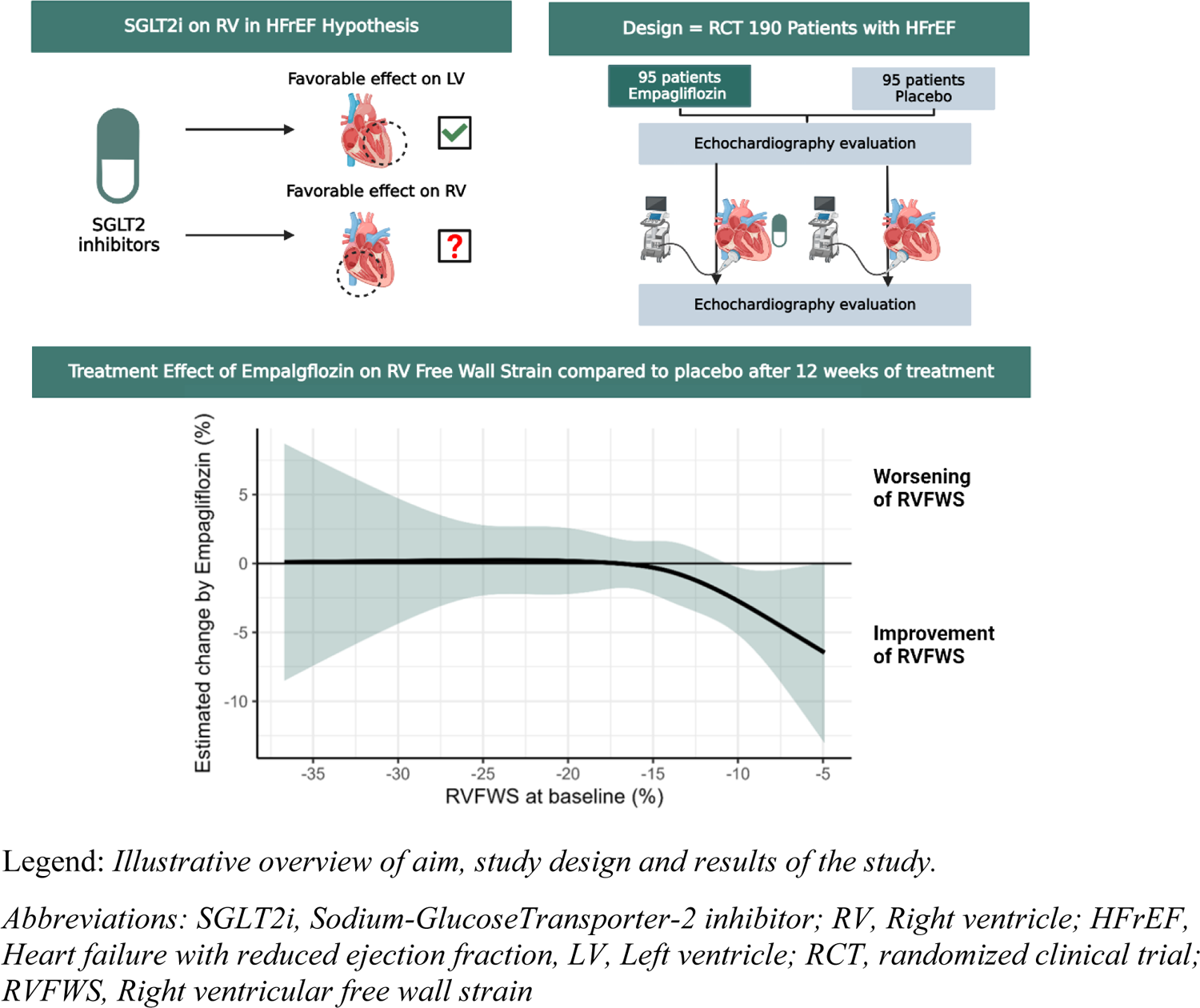

## Notes

### Competing Interest Statement

The authors have declared no competing interest.

### Clinical Trial

NCT03198585

### Author Declarations

The study was approved by the Danish National Committee on Health Research Ethics (reference no. H-17010756). All participants provided written informed consent prior to inclusion. The study was conducted in accordance with the Declaration of Helsinki and applicable national regulations. The trial is registered at ClinicalTrials.gov (Unique Identifier: NCT03198585).

